# SARS-CoV-2 Delta vaccine breakthrough transmissibility in Alachua, Florida

**DOI:** 10.1101/2021.11.10.21266134

**Authors:** Brittany Rife Magalis, Shannan Rich, Massimiliano S Tagliamonte, Carla Mavian, Melanie N Cash, Alberto Riva, Simone Marini, David Moraga Amador, Yanping Zhang, Jerne Shapiro, Amelia Horine, Petr Starostik, Maura Pieretti, Samantha Vega, Ana Paula Lacombe, Jessica Salinas, Mario Stevenson, Paul Myers, J. Glenn Morris, Michael Lauzardo, Mattia Prosperi, Marco Salemi

**Author notes:** Corresponding author Marco Salemi, Ph.D., Emerging Pathogens Institute, University of Florida College of Medicine, 2055 Mowry Rd., Gainesville, FL 32610, USA.

## Abstract

**Background:** SARS-CoV-2 Delta variant has caused a dramatic resurgence in infections in the United Sates, raising questions regarding potential transmissibility among vaccinated individuals.

**Methods:** Between October 2020 and July 2021, we sequenced 4,439 SARS-CoV-2 full genomes, 23% of all known infections in Alachua County, Florida, including 109 vaccine breakthrough cases. Univariate and multivariate regression analyses were conducted to evaluate associations between viral load (VL) level and patient characteristics. Contact tracing and phylogenetic analysis were used to investigate direct transmissions involving vaccinated individuals.

**Results:** The majority of breakthrough sequences with lineage assignment were classified as Delta variants (74.6%) and occurred, on average, about three months (104 ± 57.5 days) after full vaccination, at the same time (June-July 2021) of Delta variant exponential spread within the county. Six Delta variant transmission pairs between fully vaccinated individuals were identified through contact tracing, three of which were confirmed by phylogenetic analysis. Delta breakthroughs exhibited broad VL values during acute infection (IQR 1.2 – 8.64 Log copies/ml), on average 38% lower than matched unvaccinated patients (3.29 – 10.81 Log copies/ml, p<0.00001). Nevertheless, 49-50% of all breakthroughs, and 56-60% of Delta-infected breakthroughs exhibited VL above the transmissibility threshold (4 Log copies/ml) irrespective of time post vaccination.

**Conclusions:** Delta infection transmissibility and general VL patterns in vaccinated individuals suggest limited levels of sterilizing immunity that need to be considered by public health policies. In particular, ongoing evaluation of vaccine boosters should address whether extra vaccine doses might curb breakthrough contribution to epidemic spread.

---

According to the definition provided by the Centers for Disease Control and Prevention (CDC),^1^ several rapidly spreading variants of severe acute respiratory syndrome coronavirus 2 (SARS-CoV-2) have risen to the status of “variants of concern” (VOC), accumulating as many as 17 unique genomic mutations with respect to the original viral strain originating from the Wuhan province of China.^2^ The spread of these and other VOCs across the globe, including the United States (US), evidence of increased transmissibility, and level of evolutionary divergence from the original strain, have raised questions regarding the extent of protection of currently implemented vaccines against infection.^3-7^ Lineage B.1.1.7 (Pango nomenclature^8,9^), otherwise known as Alpha (WHO nomenclature^10^), was detected in September 2020 in Kent, UK, and was estimated to have arrived in the US approximately two months later.^11^ Other VOCs – Beta, Gamma, Delta and Epsilon – were later identified and have been circulating, at different frequencies, in different areas of the world.^3,12-15^ Lineage B.1.617 emerged in India in 2020 and diverged into three sub-lineages, known as B.1.617.1 (the ‘original’ B.1.617), B.1.617.2, renamed Delta, and B.1.617.3. ^14^ Delta carries more than a dozen mutations, including L452R, also found in the California variant Epsilon,^15^ associated with increased Spike stability and viral fusogenicity, which results in enhanced viral replication and infectivity,^16^ as well as impaired immune response through antibody neutralization.^17^

As of November 03, 2021, 66.9% of Americans were fully vaccinated,^18^ yet emergence of the Delta variant, which has rapidly been spreading in the US since June 2021, has caused a dramatic resurgence of infections and hospitalizations.^19-21^ Approximately 74% of infections with the Delta variant are followed by symptomatic onset^22^ and are characterized by high viral load (VL) linked to higher transmission rates,^23^ explaining how this variant has outpaced other VOCs and become the predominant variant in several countries, mostly driven by dissemination among the unvaccinated. ^24^

According to clinical trial results, a two-dose regimen of Pfizer-BioNTech (BNT162b2) vaccine confers 94.6% protection against severe disease,^6^ while the Moderna (mRNA-1273) vaccine confers 94.1%.^25^. While vaccination continues to provide excellent protection against hospitalization and death, over the past six months there has been a gradual decline in vaccine efficacy against infection.^26^ In the US, these vaccine breakthrough cases linked to VOCs have been reported since January 2021.^27^ In July 2021, following multiple large public events in a Barnstable County, Massachusetts, town, an outbreak (90% Delta) of infection (albeit mild disease) was identified among Massachusetts residents traveling recently to the town, primarily comprised of fully vaccinated persons.^19^ Within the same month, a similar report of widespread Delta circulation (95% of sequenced samples by July 24) among individuals in Wisconsin (USA) was published in medRxiv.^28^ Both studies reported similar VL among vaccinated and unvaccinated individuals, which were estimated by using the number of polymerase chain reaction cycles required to quantify viral genetic sequence fragments (CT value) in a specimen. Previous work has shown that infectious SARS-CoV-2 can usually be recovered from specimens with CT < 25 – 30.^29^ Yet, CT values are only a proxy for the level of virus shed in the nasal passages and, while these data suggest vaccinated individuals are susceptible to infection by the Delta variant and harbor the level of infectious virus required for further transmission, definitive evidence of this transmission has not been presented. Moreover, CT values do not allow for a quantitative evaluation of the VL threshold required for potential transmission within a specific population, e.g., the fraction of vaccine breakthrough cases harboring a VL compatible with secondary transmissions, which both empirical data and theoretical studies have shown to be 4 Log copies/ml.^30,31^ Finally, a recent comparison of Moderna (mRNA-1273) and Pfizer/BioNTech (BNT162b2) vaccines in cohorts from states with high prevalence of Delta infections (Minnesota, Wisconsin, Arizona, Florida, and Iowa) showed strong protection against disease but also a lower risk of infection after full vaccination with mRNA-1273 than after full vaccination with BNT162b2,^32^ although the study did not provide direct virus genomic data of the breakthrough cases.

The present study analyzes data generated as part of the SARS-CoV-2 genomic epidemiology surveillance program in Alachua County, Florida, from October 2020 to August 2021, to answer three main questions: 1) Did the emergence of the Delta variant result in increases in vaccine breakthroughsã 2) Do fully vaccinated people, infected by the Delta variant (or other variants), transmit the infectionã 3) What fraction of vaccine breakthrough patients have a VL above the transmissibility threshold during acute infection?

## METHODS

### SARS-COV-2 genomic epidemiology surveillance in Alachua County, Florida

The Alachua County Department of Health, with assistance from designated UF personnel, has been responsible for contact tracing efforts for University of Florida (UF) students, faculty, staff, and other UF-affiliated people, including the UF Health Academic Medical Center (∼123,000 total UF Affiliates) since the beginning of the epidemic. SARS-CoV-2 positive samples were collected for virus full genome sequencing, as part of this program, between October 2020 and August 2021 from patients hospitalized at UFHealth Shands Hospital Gainesville during this time period, as well as from testing sites of UF PathLabs in Gainesville serving Alachua County residents (Supplementary Table S1). Samples from positive patients in the Tampa Bay are and Miami Dade (provided by BayCare and University of Miami, respectively), which had been collected for other studies, were included in the sequence analysis for comparison purposes (Table S1). Full epidemiological investigations were conducted on Alachua County positive cases to collect exposure information, trace contacts, and provide disease transmission education.

### Vaccine breakthrough cases involvement

Vaccine breakthrough cases were defined as individuals who were PCR-positive for SARS-CoV-2 and≥14 days after the second dose of Pfizer or Moderna or first dose of Janssen/J&J. Repeat saliva samples were collected from possible breakthrough cases for sequencing. Patient samples and linked data were fully deidentified before sample processing. The study was reviewed and approved under the category of Public Health practice by the University of Florida Institutional Review Board (IRB) and the Florida Department of Health IRB.

### Sample processing and next generation sequencing

Viral RNA, from either 200 μL of viral transport medium for NP swabs or 180 μL of saliva, was extracted for each sample using the QIAamp 96 Viral RNA Kit with the QIAcube HT (Qiagen, Germantown, MD) using the following settings with a filter plate: the lysed sample was premixed 8 times before subjecting to vacuum for 5 minutes at 25kP and vacuum for 3 min at 70kPa. Following 3 washes using the same vacuum conditions above, the samples were eluted in 100 μL AVE buffer followed by a final vacuum for 6 minutes at 60kPa. Nine microliters of RNA were used for cDNA synthesis and library preparation using the COVIDSeq Test kit (Illumina, San Diego, CA) and Mosquito HV Genomics Liquid Handler (SPT Labtech Inc., Boston MA). The size and purity of the library was determined using the 4200 TapeStation System (Agilent, Santa Clara, CA) and the Qubit dsDNA HS Assay Kit (Life Technologies, Carlsbad, CA) according to the manufacturer’s instructions. Constructed libraries were pooled and sequenced using the NovaSeq 6000 Sequencing System SP Reagent Kit and the NovaSeq Xp 2-Lane Kit. Illumina’s DRAGEN pipeline was used to derive sample consensus sequences, which were filtered based on a minimum of 70% coverage of the genome and 20X sequencing depth.

### Database sequence retrieval, sequence alignment, and masking

For each in-house sequencing run, sequences within the GISAID database associated with the state of Florida were extracted up to August 03, 2021, and added to the collection of high-coverage in-house-produced genome sequences (N=3,110 from Alachua County, N=126 from Fort Myers, N=791, from Miami Dade), totaling 20,117 sequences, ranging from February 28, 2020, to August 03, 2021. Each Floridian sequence within this concatenated dataset was then used in a local alignment (BLAST)^33^ search for the most (genetically) similar non-Floridian sequence in the GISAID database as of the current date and linked to two reference sequences including the best match (highest E-value) with a date occurring within one month following, as well as one month prior to, the sampling date of the Floridian sequence.^34^ After removing duplicate sequences (sequences with same GISAID ID), the final GISAID reference dataset totaled 5,074, ranging from February 21, 2020, to July 27, 2021. Sequences were aligned in viralMSA^35 35^ using the MN908947 reference sequence. Mutations potentially associated with contamination, recurrent sequencing errors, or hypermutability were masked using a VCF filter provided at https://virological.org/t/masking-strategies-for-sars-cov-2-alignments/480.

### Phylogenetic tree reconstruction and optimization

Following the first in-house sequencing run and first round of BLAST protocol described above, the multiple sequence alignment for all sequences up to November 25, 2020, was used in the reconstruction of an initial maximum likelihood tree in IQ-TREE2 using the best-fit evolutionary model according to Bayesian Information Criterion ^36^.^36^. The tree was updated with each round of sequencing, concatenation, and local alignment using phylogenetic placement in UShER.^37^ The tree topology was also refined after each round using two rounds of subtree pruning and regrafting (maximum length of each move of 100) in FastTreeMP.^38^. Branch lengths were re-optimized using the gamma distribution. Lineages for all sequences were determined using the PangoLEARN model (updated daily), which is trained using GISAID SARS-CoV-2 sequences to classify incoming sequences based on molecular and epidemiological criteria.^8,9^

### SARS-CoV-2 Delta transmission cluster identification

The subtree containing sequences classified as Delta was pruned from the full tree for transmission cluster analysis. Root-to-tip regression was used to identify outliers, defined as sequences with extreme deviation in genetic divergence from the best-fit root of the tree, suggestive of incorrectly reported sampling dates, though true variation owing to natural processes such as recombination and positive selection cannot be ruled out.^39^. Two sequences were identified as outliers and pruned from the tree prior to downstream analysis using the ape package in R.^40^ Transmission cluster identification within the resulting pruned tree was performed using a depth-first search among nodes restricted to minimal evolution (branch lengths) and sampling time difference (available from https://github.com/ProsperiLab/Dynamite). This transmission cluster identification approach is similar in nature to other phylogenetic cluster-picking algorithms (e.g., Phylopart^41^Phylopart^41^, ClusterPicker^42^), with the exception that clusters were not restricted to monophyletic clades and an informed serial time interval was used to accept or reject sampled sequences based on a reasonable transmission window. As with other phylogenetic algorithms, a genetic distance threshold was required as criterion to accept or reject nodes into each cluster. Similar to Phylopart, the optimal threshold was chosen which maximized the number of clusters identified-4.23 × 10^−5^ substitutions/site. Phylopart was also used for robust comparison, which resulted in one less cluster than the described algorithm for a range of optimum thresholds (3.86 × 10^−5^ – 5.03 × 10^−5^). For the described depth-first algorithm, individual nodes (either external or internal) were accepted as part of a cluster if the mean branch length of this node and all preceding accepted nodes did not exceed the optimal threshold. The serial time interval used as an additional criterion to accept tips into individual clusters was 6 days, based on meta-analysis performed by Rai *et al*. for SARS-CoV-2.^43^ Similar to the branch length procedure, external nodes were accepted as part of a cluster if the above criterion was met and if the mean pairwise sampling time difference among the tip in question and previously accepted tips did not exceed the specified serial time interval threshold.

### Viral load quantification

Levels of SARS-CoV-2 were determined using the 2019-nCoV_N1 assay (primer and probe set) with 2019-nCoV_N_positive control (IDT, Coralville, Iowa). Viral RNA was extracted as previously described then subjected to first strand synthesis using ProtoScript II Reverse Transcriptase according to the manufacturer’s instructions (NEB, Ipswich, MA). Quantitative PCR was performed using TaqMan Fast Advanced Master Mix (Waltham, MA) according to the manufacturer’s instructions. A standard curve was generated using N1 quantitative standards 10-fold diluted to determine viral copies. The assay was run in triplicate including one non-template control.

### Relationship between VL, vaccination status and SARS-CoV-2 lineage

We performed a linear regression on the VL as dependent variable, with vaccination status (yes/no), Delta lineage (yes/no, age (<21, 21-28, 28-44, >44 years old or unknown), gender (male, female, unknown), and days from January 2021 as covariates. We also considered an interaction term between vaccination status and Delta, and coefficients were calculated with and without the inclusion of the interaction term. For interpretation of individual coefficients, we can assume that either vaccination status or Delta variant might be confounded by age, gender and time passed, and give a possibly causal interpretation of said coefficients as total effects under that adjustment set. Univariable and multiple regression analyses were performed to evaluate associations between patient characteristics and either VL or RT-PCR cycle threshold (CT) levels. The analyses considered demographic variables [age (years), sex (male vs. female), race (white vs. non-white), ethnicity (Hispanic vs. non-Hispanic)] and clinical information [vaccine brand (Moderna, Pfizer, or J&J), symptom status (yes/no)], and time between vaccination and onset (days). To investigate associations between these variables and levels of infectivity, outcomes were also categorized as binary variables for multivariable logistic regression analysis. Patients were considered above the transmissibility threshold (i.e., potentially infectious) if they had a VL level greater than or equivalent to Log 4 copies per ml of sample. ^31^

## RESULTS

Between October, 2020, and first week of August, 2021, we sequenced 4,439 SARS-CoV-2 samples from patients in Alachua County (Table S1), Florida, representing approximately 22% of reported positive cases during the same period (20,612 cases between October 05, 2020, and August 06, 2021). Following a trend similar to the rest of the US, the first VOC circulating in the county, Alpha, was detected in December 2020. Alpha became the dominant variant in March 2021 but decreased in favor of VOC Gamma and other B.1 sub-variants in the following months (Figure 1A), while the number of infections significantly dwindled due in part to a major vaccination effort,^18^ which began in Alachua County in December 2020. Sporadic vaccine breakthroughs were identified in the first seven months of 2021 (Figure 1B), including five Alpha variant cases, two Epsilon, five B.1, three B.1.2, one B.1.596, and one B.1.377. During the third week of June, we detected the first Delta infection in a fully vaccinated patient, followed by a rapid increase in Delta vaccine breakthroughs through the end of July, coinciding with Delta becoming the most prevalent variant (Figure 1A and 1B). Overall, 109 vaccine breakthrough cases were identified, including 58 Delta infections and 34 unknowns due to low coverage of the Spike protein. The breakthrough cases exhibited an average time between full vaccination (defined as 14 days following final dose) and COVID-19 diagnosis of about 3 months (mean = 101.6 ± 57.7 days) (Figure 1C). Compared to the number of vaccinations, the proportion of diagnosed breakthrough infections remained extremely low throughout our surveillance (Supplementary Figure S1), in line with several other studies that have shown the effectiveness of currently available vaccines. However, while the spike in breakthrough cases during the month of July did coincide with a significant vaccination scale up in Alachua County^18^, the majority (71%) of these patients had already been fully vaccinated for more than 90 days (Figure 1C) and became infected by the Delta variant in July, while its frequency was exponentially increasing among the unvaccinated population (Figure 1B), indicating that the increase in breakthroughs was indeed linked with the emergence of the Delta variant.

**Figure 1.**
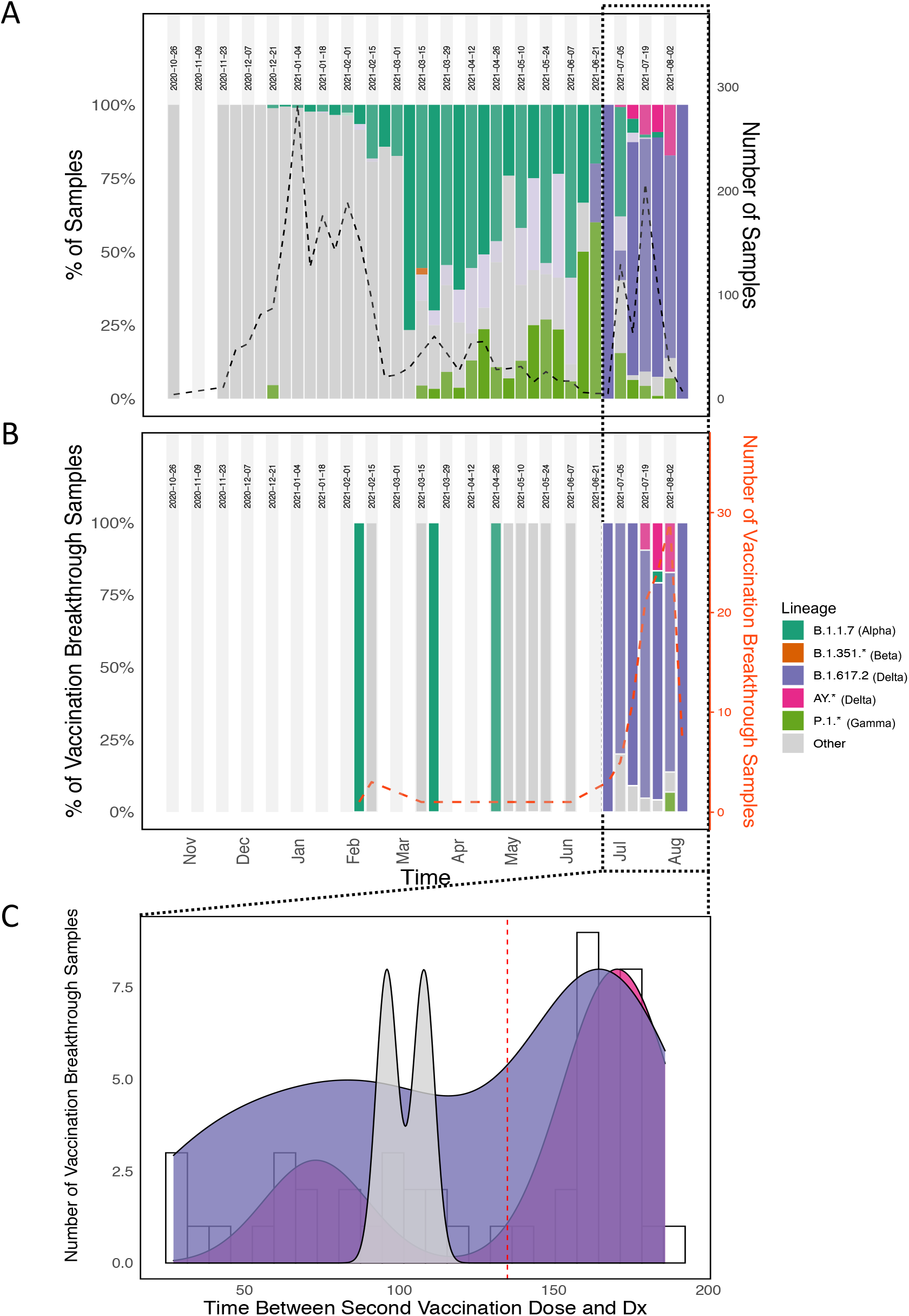
SARS-CoV-2 variants distribution in Alachua County, Florida, over time. (A) Lineage distribution (y-axis) *vs*. time (x-axis) among sequenced samples. Total number of samples successfully sequenced is represented by the black, dotted line (right y-axis). (B) Lineage distribution (y-axis) *vs*. time in vaccine breakthrough cases. Total number of samples successfully sequenced is represented by the red, dotted line (right y-axis). (C) number of vaccine breakthrough cases (x-axis) vs. time between 2^nd^ vaccination dose and diagnosis (y-axis).

The breakthrough population consisted largely of White individuals (73.4%), ages 36.7 years on average (SD=14.2), all of whom had mild symptoms with low prevalence of associated comorbidities (Table 1). None of the individuals within this population required hospitalization. Approximately 44% of breakthrough cases could be traced to known exposures, identifying household transmission (54.2%) as the primary source of putative infection, followed by community (37.5%) and healthcare (6.3%) related exposures. A supplemental saliva sample was collected from a subset of the breakthrough cases (N=83), on average 4.2 days after disease onset (Table 1), to measure the VL during acute infection. VL among breakthrough cases infected with the Delta variant (N=56) averaged 4.66 Log copies/ml, with an interquartile range (IQR) of 1.2 – 10.62, overlapping to the one observed among breakthroughs infected by other variants (N=13) with average VL of 5.39 Log copies/ml, and IQR 1.41 – 8.36 (*p* = 0.35 from a two tailed Mann–Whitney U test). For comparison with the non-vaccinated population, VL was also evaluated in age- and gender-matched data sets, retrospectively assembled for the months of January – April and July, that included randomly selected independent samples (i.e., samples from patients not directly linked through known transmission events) from non-vaccinated individuals infected with Delta (N=36) or other variants (N=75). In agreement with previous reports^34^, unvaccinated patients infected with the Delta variant exhibited, on average, the highest VL (mean 7.36 Log copies/ml, IQR 3.29 – 10.81) compared to vaccinated Delta or non-Delta breakthroughs (Figure 2), as well as to unvaccinated patients infected by other variants (mean 6.15 Log copies/ml, IQR 3.56 – 10.92), although effect size was modest (6% increase, *p* = 0.17 from a two tailed Mann–Whitney U test), probably due to small sampling size. Contrary to other reports^19,28^, however, Delta-infected breakthrough cases in Alachua County exhibited an average 38% VL reduction compared to unvaccinated Delta cases (*p*<0.00001, two tailed U-test, after Bonferroni correction), and 34% (*p*<0.00001) compared to unvaccinated non-Delta cases (Figure 2). The multivariable analysis did not find any association between VL and age, gender, race, ethnicity, or vaccine type in the sample (Table S2). Yet, Delta infections had a strong association with VL above the transmissibility threshold of Log 4 RNA copies/ml,^30,31^ with a 2.46 odd ratio (OR) and confidence interval (CI) 1.05 – 5.97 in the unadjusted analysis, and OR 3.04 and CI 1.16 – 8.54 in the adjusted analysis (Table S3). In particular, the majority of vaccine breakthrough cases infected with the Delta variant (58.5%) exhibited a VL above the required threshold for potential transmission (Figure 2).

**Table 1.** Summary of vaccine breakthrough population (N=109) in Alachua County, Florida from January to August 2021. *Numbers are present as frequencies and (percentages) unless otherwise stated*

|  |  |
| --- | --- |
| **Age (years)** | 36.7 (14.2) |
| **Sex** |  |
| Female | 68 (62.4%) |
| Male | 41 (37.6%) |
| **Race** |  |
| White | 80 (73.4%) |
| African American/Black | 10 (9.2%) |
| Asian/Pacific Islander | 11 (10.1%) |
| Other/Unknown | 8 (7.3%) |
| **Ethnicity** |  |
| Non-Hispanic | 91 (83.5%) |
| Hispanic | 18 (16.5%) |
| **Symptoms** | 96 (88.1%) |
| Dry cough | 47 (43.1%) |
| Productive cough | 18 (16.5%) |
| Dyspnea | 14 (12.8%) |
| Anosmia | 35 (32.1%) |
| Ageusia | 34 (31.2%) |
| Sore throat | 40 (36.7%) |
| Headache | 53 (48.6%) |
| Runny nose | 79 (72.5%) |
| Fatigue | 54 (49.5%) |
| **Comorbidities** |  |
| Asthma | 11 (10.1%) |
| Diabetes | 3 (2.8%) |
| Hypertension | 15 (13.8%) |
| BMI <sup>b</sup> | Mean = 27.0 (SD = 6.3) |
| **Known exposure (Y)** | 48 (44.0%) |
| Household | 26 (54.2%) |
| Community | 18 (37.5%) |
| Healthcare | 3 (6.3%) |
| **Vaccine** |  |
| BNT162b2 (“Pfizer/BioNTech”) | 83 (76.1%) |
| mRNA-1273 (“Moderna”) | 11 (10.1%) |
| Ad.26.COV2.S (“Johnson & Johnson/Janssen”) | 14 (12.8%) |
| NVX-CoV2373 (“Novavax”) | 1 (0.92%) |
| Time between vaccination and disease onset <sup>a</sup> (days) | Mean = 104.0 (SD = 57.5) |
| Time between disease onset <sup>a</sup>, and sample collection date (days) for VL measurement (N=83) | Mean = 4.2 (SD = 2.4) |

<sup>a</sup> Disease onset is defined as the date of symptom onset for symptomatic individuals or the original date of lab collection for asymptomatic individuals
<sup>b</sup> BMI: body mass index.

**Figure 2.**
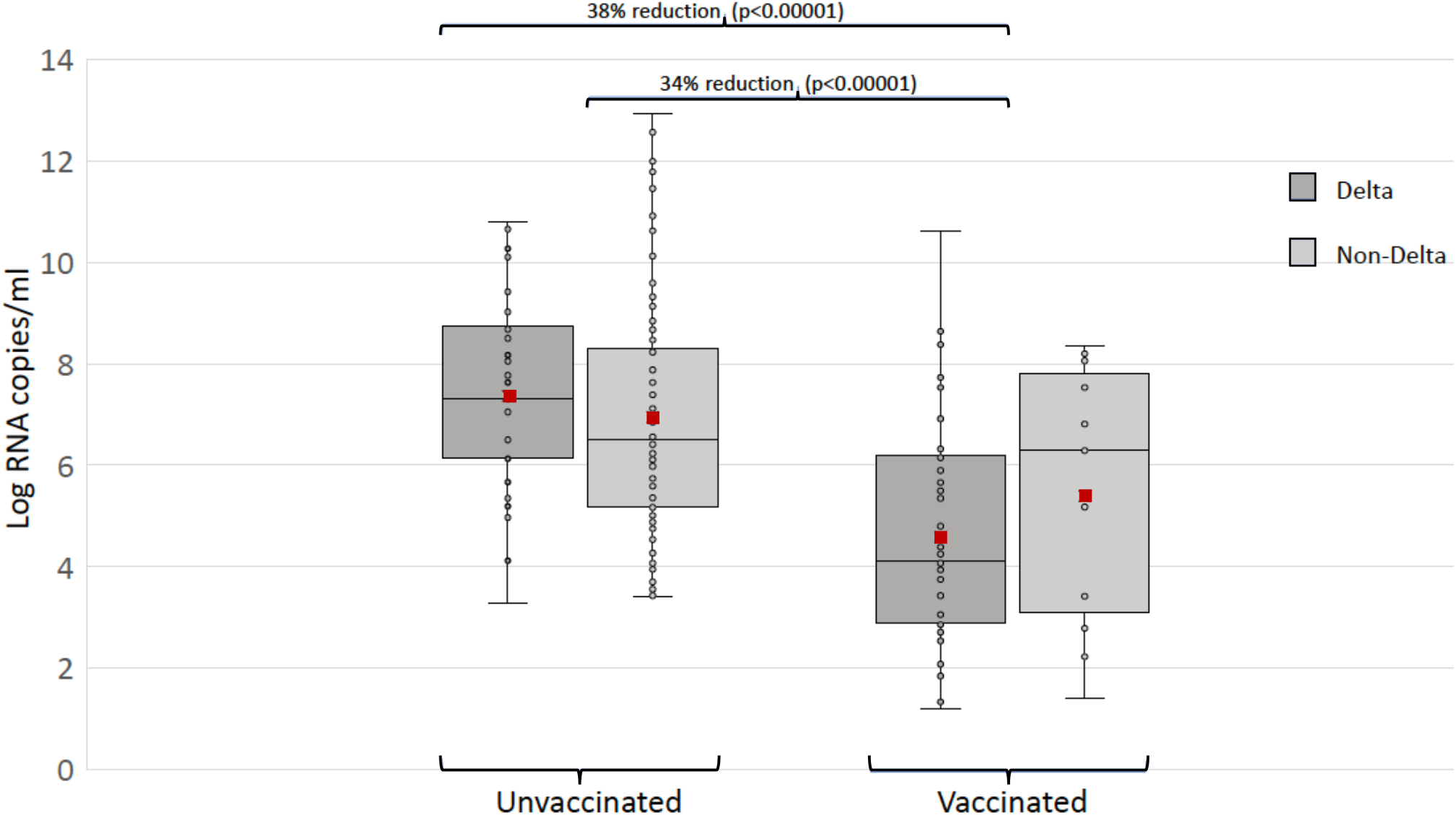
VL distribution in vaccinated and unvaccinated SARS-CoV-2 infected patients. Each box plot (with line at median and red dot indicating the mean) represents a group of patients infected with either the Delta variant or other variants, vaccinated or unvaccinated according to the legend in the figure. For each group pair, a two-tailed Mann–Whitney U test was executed. P-value and effect size are shown on top for those comparisons between groups that were significant at the 5% level after Bonferroni correction for multiple tests.

Direct virus transmission among vaccinated individuals infected by the Delta variant was, indeed, confirmed by contact tracing and phylogenetic analysis of SARS-CoV-2 sequence data. Contact tracing allowed us to identify six putative transmission pairs, each one involving a fully vaccinated Donor (D), part of our initially detected breakthrough cases, and a Recipient (R) with no other known infection exposure history (Table 2). At the time of symptom onset, Donor D1 had been fully vaccinated with mRNA-1273, for 120 days, while Donors D2 – D6 with BNT162b2 for 143 – 176 days, with all of them exhibiting VL > Log 4 RNA copies/ml. Four out of six Recipients had also been vaccinated with BNT162b2 for 67 – 164 days at the time of symptom onset, thus classifying them as additional vaccine breakthrough cases. For three of these D-R pairs, we were able to obtain saliva samples collected approximately four days following initial diagnosis and obtain the full genome sequence of the virus. Sequences for individuals involved in transmission pairs (all Delta variants) were evaluated in the context of Delta sequences derived from other parts of Florida and of closely related Delta sequences retrieved from the GISAID database (see Methods). Four non-monophyletic transmission clusters were identified using a depth-first search algorithm applied to the phylogeny, identified as groups of individuals sharing minimal genetic sequence differences and pairwise sampling time differences within the reported typical serial time interval of up to six days (see Methods). Though geographical sampling locations were intermixed throughout the phylogeny, clusters were primarily comprised of local transmissions (Figure 3A). Vaccination breakthroughs were also intermixed throughout the phylogeny; however, all contact tracing-identified transmission pairs belonged to cluster c3 (Figure 3B). Each receiving (R) individual within a transmission pair was observed directly adjacent to the corresponding putative donor (D) individual, validating direct transmission between pairs. All six individuals belonging to the three successfully sequenced transmission pairs were fully vaccinated, supporting the transmission capability of fully vaccinated individuals.

**Table 2.** Vaccine and Viral load information for identified transmission pairs.

| **Pair** | **Individual** | **Vaccine** | **Days since vaccination\*** | **VL** |
| --- | --- | --- | --- | --- |
| D1-R1 | **D1** | Moderna | 120 | **4.53** |
|  | R1 | Pfizer | 139 | 2.08 |
| D2-R2 | **D2** | Pfizer | 162 | **6.81** |
|  | R2 | Pfizer | 164 | NA |
| D3-R3 | **D3** | Pfizer | 143 | **8.64** |
|  | R3 | Pfizer | 67 | 3.94 |
| D4-R4 | **D4** | Pfizer | 157 | **4.12** |
|  | R4 (Uncollected) | NA | NA | NA |
| D5-R5 | **D5** | Pfizer | 173 | **4.07** |
|  | R5 (No sequence) | Pfizer | 86 | NA |
| D6-R6 | D6 | Pfizer | 176 | NA |
|  | R6 (Uncollected) | NA | NA | NA |

\* Time since vaccination was defined as two weeks after administration of 2<sup>nd</sup> dose of either Moderna or Pfizer vaccine.

**Figure 3.**
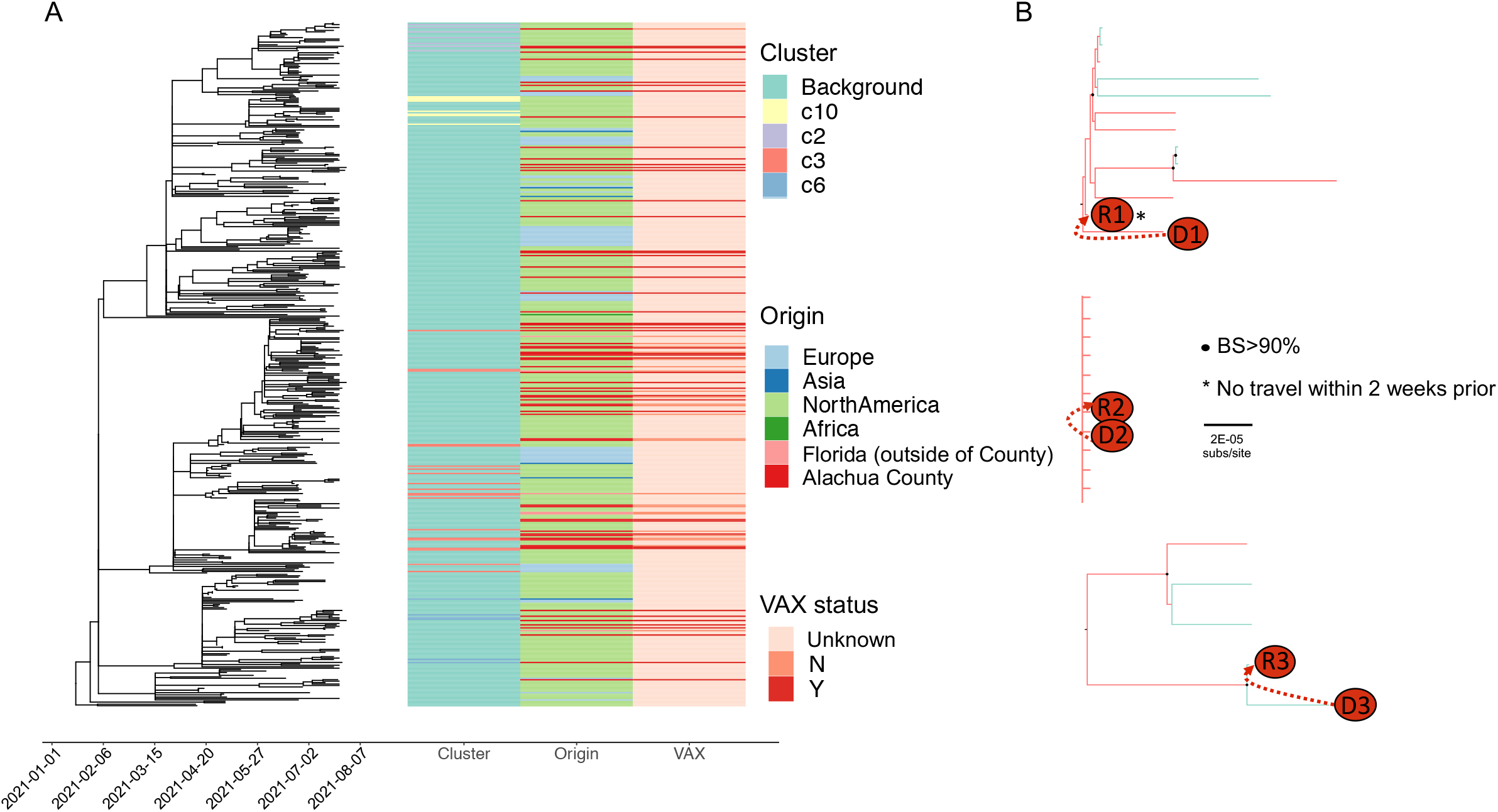
Phylogenetic reconstruction and transmission characterization of SARS-CoV-2 Delta sequences obtained from Alachua County, Florida, and epidemiologically relevant outside locations. (A) Phylogeny of sequences with heatmap depicting cluster origin, geographical origin, and vaccination status of each sampled sequence. (B) Phylogenetic relationships of sequences from donor(D)-recipient(R) pairs linked via exposure using contact tracing (branches are colored according to cluster origin). Bootstrap support (BS) within clades containing transmission pairs >90% are represented by black dots. Individuals with confirmed no known travel within 2 weeks prior to diagnosis are represented with asterisks. Branches are scaled in substitutions/site.

To examine further SARS-CoV-2 Delta variant potential transmissibility in breakthrough infections in the context of the specific vaccination received by the patients, we investigated the correlation between VL and time since full vaccination in individuals who received the Pfizer-BioNTech (N=72 assigned variants, N=53), the Johnson & Johnson/Janssen (N=9, N=6 Delta), or the Moderna (N=8, N=6 Delta) vaccine. The only patient in our population sample vaccinated with Novavax was excluded. As regression analyses assume random sampling of the population, individuals included in transmission clusters are considered to be potentially linked epidemiologically and should not be considered independent. Minimal branch support within cluster c3 could not rule out epidemiological linkage among the three D-R transmission pairs (Figure 3). Therefore, the three donor individuals were removed prior to regression analysis, leaving the three recipient individuals which we know to be unrelated via direct transmission. Remaining Delta vaccination breakthroughs with VL data were sparsely placed throughout the tree (45 in background population and remaining two in clusters c2 and c6) and could thus be included. Only one unvaccinated Delta individual with VL data was observed within a transmission cluster (c2) and was considered to be distinct from the vaccinated individual in this cluster by significant branch support and so was included in regression analysis. Non-Delta sequences were not included in the phylogeny; however, samples from this population were chosen for VL analysis so that a wide range of collection times and variants were included, minimizing the probability of epidemiological linkage and the need for phylogenetic assessment of independence. Multiple regression analysis had already found no evidence of association between VL and time since full vaccination (8 – 186 days), time interval between symptom onset and sampling, or vaccine type using either linear or logistic methods (Supplementary Table S2). There was also no correlation between distribution of VL over time elapsed since full vaccination, defined as the time interval between two weeks after 2^nd^ vaccination dose (in case of Pfizer-BioNTech or Moderna) or after single dose (Johnson & Johnson/Janssen). Importantly, the proportion of patients with VL above the theoretical transmission threshold (VL > 4 Log copies/ml) was essentially the same when looking at patients infected <100 days (48.8% all, 55.6% Delta) or >100 days (50% all, 60% Delta) after full vaccination (Figure 4 The analysis gave similar results when only Delta vaccine breakthrough patients vaccinated with Pfizer (N=42) were considered (Supplementary Figure S1). Given the small number of Delta vaccine breakthrough patients who received Johnson & Johnson/Janssen (N=6) or Moderna (N=6), no conclusions could be drawn, although the few Moderna vaccinated patients were the only ones who showed a strong linear correlation (R^2^ = 0.86) between VL, and time elapsed since full vaccination (Supplementary Figure S2).

**Figure 4.**
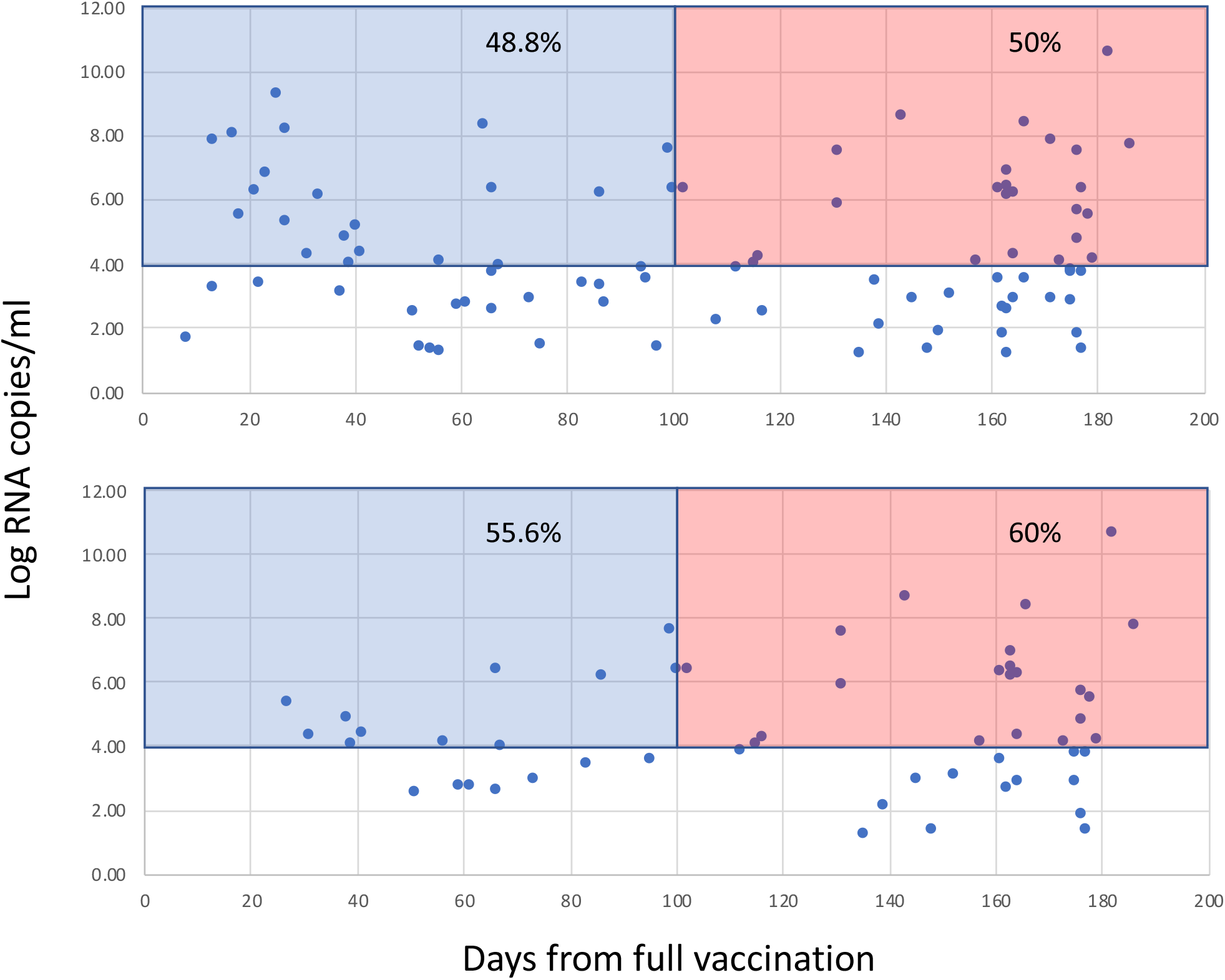
VL vs. time from full vaccination in vaccine breakthrough cases at the time of symptoms onset. Each dot in the scatterplots represents a single patient fully vaccinated with either Pfizer/BNT162b2, Moderna/mRNA-1273 or Johnson & Johnson/Janssen vaccine. The Y-axis reports the VL measure in Log RNA copies/ml. The x-axis represents the time (in days) between full vaccination, defined as two weeks after 2^nd^ vaccination dose (in case of Pfizer-BioNTech or Moderna) or after single dose (Johnson & Johnson/Janssen), and time of sampling, which occurred for each patient on average 4.2 days after symptoms onset (see Table 1). Shaded areas highlight proportions patients (reported inside each area) with VL above the transmissibility threshold (Log 4 copies/ml) who were full vaccinated for <101 days (cyan) or >100 (pink). The top panel includes vaccine breakthrough cases infected with different SARS-CoV-2 variants (N=89). The bottom panel includes only patients infected with the Delta variant (N=53).

## DISCUSSION

The rapid emergence, in July 2021, of Delta infections correlated with a major spike in reported cases, hospitalizations, and deaths in Florida and in Alachua County, due primarily to infections in unvaccinated persons^44^. It also coincided with a sudden spike in vaccine breakthrough cases, demonstrating that direct transmission from vaccine breakthroughs can occur, with more than half of the Delta breakthroughs in our sample harboring sufficient viral copies to transmit the virus during acute infection.

Overall, the number of breakthrough cases detected, during our nearly nine-month genomic epidemiology surveillance program, represented approximately 2% of the sequenced cases, a low number of vaccine failures in agreement with the known effectiveness of the vaccines according to randomized clinical trials. Even assuming that we may have missed a substantial number of vaccine breakthrough infections in asymptomatic individuals, we have no records of severe/fatal COVID-19 cases that involved vaccinated patients between December 2020 (when vaccination began in Alachua County) and end of July 2021. Our VL data also show that, despite Delta variant high transmissibility – as evidenced by its ability to become the majority variant within a month since its emergence in the county – vaccination is associated with lower viral replication in breakthrough cases compared to unvaccinated patients. While other studies have reported similar VL among vaccinated and unvaccinated individuals^19,28^, such studies utilized CT values as a proxy for VL among groups of patients rather than the actual number of RNA copies, which may in part explain the discrepancy. Moreover, studies such as the one in Barnstable County^19^, Massachusetts, focused on outbreaks within local communities likely characterized by related transmission clusters that may confound statistical comparisons under the assumption of independent sampling (because samples from different patients can be related through a transmission chain), unless adjusted by using phylogenetic regression methods ^45,46^. Unfortunately, none of those studies investigated the phylogenetic relationships among the samples to exclude potentially linked individuals. Our VL analysis, on the other hand, was filtered on individuals that were considered independent transmission events through careful sampling and transmission cluster identification within the phylogeny.

Development of sterilizing immunity does not commonly occur for most human and animal vaccines, and current vaccine trials are not designed to address it since only clinically affected subjects are usually tested for the virus. Nevertheless, assessing SARS-CoV-2 transmissibility in vaccine breakthrough cases is crucial, especially in light of the current spread of the Delta variant, not only to break the cycle of viral transmission, thus resulting in fewer cases of severe COVID-19 and death, but also to reduce the likelihood of emergence of more pathogenic or potentially vaccine-resistant viral variants. The continuous circulation of SARS-CoV-2 among unvaccinated individuals, as well as vaccinated who, albeit protected from severe disease, are susceptible from infection and can potentially contribute to further transmission, provides the virus the chance to continue exploring the fitness landscape and accumulating mutations that may eventually result in the emergence of even more transmissible/pathogenic or vaccine-resistant strains. Therefore, while additional observations based on larger numbers of patients are necessary, our work indicate that the differential level of sterilizing immunity, or lack of thereof, that may be present in the vaccinated population, is an important factor to be considered for the implementation of the next phase of vaccination and intervention policies. In particular, the ongoing evaluation of vaccine boosters^47,48^ should address, besides effectiveness against disease, whether the administration of extra vaccine doses may reduce the number of breakthrough cases, or at least decrease the proportion of breakthroughs exhibiting VL above the transmissibility threshold during the acute phase of infection, thus improving sterilizing immunity.

## Data Availability

All data produced in the present study are available upon reasonable request to the authors

## Acknowledgments

This work was supported by the Stephany W. Holloway University of Florida Chair and by funds of the University of Florida Office of Research and Health Science Center with resources from the Interdisciplinary Center for Biotechnology Research Gene Expression Core (RRID:SCR_019145), NextGen Sequencing Core (RRID:SCR_019152) and Bioinformatics Core (RRID:SCR_019120). Funding for this work was also provided by National Science Foundation (NSF) – Division of Environmental Biology (DEB) award no. 2028221, The AIDS HealthCare Foundation Award No. OS00000633 and The Rockefeller Foundation Award 2021 HTH 012. The co-authors are thankful to Maureen Long for her critical reading of the manuscript.

## References

1. CDC. SARS-CoV-2 Variant Classifications and Definitions. (https://www.cdc.gov/coronavirus/2019-ncov/variants/variant-info.html).

2. Rambaut A, Loman N, Pybus O, et al. Preliminary genomic characterisation of an emergent SARS-CoV-2 lineage in the UK defined by a novel set of spike mutations. Virologicalorg 2020;https://virological.org/t/preliminary-genomic-characterisation-of-an-emergent-sars-cov-2-lineage-in-the-uk-defined-by-a-novel-set-of-spike-mutations/563.

3. Madhi SA, Baillie V, Cutland CL, et al. Efficacy of the ChAdOx1 nCoV-19 Covid-19 Vaccine against the B.1.351 Variant. N Engl J Med 2021 (In eng). DOI: 10.1056/NEJMoa2102214.

4. Buss LF, Prete CA, Abrahim CMM, et al. Three-quarters attack rate of SARS-CoV-2 in the Brazilian Amazon during a largely unmitigated epidemic. Science 2021;371(6526):288–292. (In eng). DOI: 10.1126/science.abe9728.

5. Sabino BD, Alonso FOM, Oliveira MSC, Venceslau MT, Guimarães MAAM, Varella RB. Long-term intermittent detection of SARS CoV 2 in the upper respiratory tract: what is the meaning of itã Infect Dis (Lond) 2021;53(2):151–153. (In eng). DOI: 10.1080/23744235.2020.1837944.

6. Polack FP, Thomas SJ, Kitchin N, et al. Safety and Efficacy of the BNT162b2 mRNA Covid-19 Vaccine. N Engl J Med 2020;383(27):2603–2615. (In eng). DOI: 10.1056/NEJMoa2034577.

7. Lopez Bernal J, Andrews N, Gower C, et al. Effectiveness of Covid-19 Vaccines against the B.1.617.2 (Delta) Variant. N Engl J Med 2021;385(7):585–594. (In eng). DOI: 10.1056/NEJMoa2108891.

8. Rambaut A, Holmes EC, O’Toole Á, et al. A dynamic nomenclature proposal for SARS-CoV-2 lineages to assist genomic epidemiology. Nat Microbiol 2020;5(11):1403–1407. (In eng). DOI: 10.1038/s41564-020-0770-5.

9. Rambaut A, Holmes EC, O’Toole Á, et al. Addendum: A dynamic nomenclature proposal for SARS-CoV-2 lineages to assist genomic epidemiology. Nat Microbiol 2021;6(3):415. (In eng). DOI: 10.1038/s41564-021-00872-5.

10. WHO. Tracking SARS-CoV-2 Variants. (https://www.who.int/en/activities/tracking-SARS-CoV-2-variants/).

11. England PH. SARS-CoV-2 variants of concern and variants under investigation in England. Technical briefing 23. 2021.

12. Tegally H, Wilkinson E, Giovanetti M, et al. Detection of a SARS-CoV-2 variant of concern in South Africa. Nature 2021;592(7854):438–443. (In eng). DOI: 10.1038/s41586-021-03402-9.

13. Tagliamonte MS, Mavian C, Zainabadi K, et al. Rapid emergence and spread of SARS-CoV-2 gamma (P.1) variant in Haiti. Clin Infect Dis 2021 (In eng). DOI: 10.1093/cid/ciab736.

14. Adam D. What scientists know about new, fast-spreading coronavirus variants. Nature 2021;594(7861):19–20. (In eng). DOI: 10.1038/d41586-021-01390-4.

15. Zhang W, Davis BD, Chen SS, Sincuir Martinez JM, Plummer JT, Vail E. Emergence of a Novel SARS-CoV-2 Variant in Southern California. JAMA 2021;325(13):1324–1326. (In eng). DOI: 10.1001/jama.2021.1612.

16. Motozono C, Toyoda M, Zahradnik J, et al. SARS-CoV-2 spike L452R variant evades cellular immunity and increases infectivity. Cell Host Microbe 2021;29(7):1124–1136.e11. (In eng). DOI: 10.1016/j.chom.2021.06.006.

17. Starr TN, Greaney AJ, Dingens AS, Bloom JD. Complete map of SARS-CoV-2 RBD mutations that escape the monoclonal antibody LY-CoV555 and its cocktail with LY-CoV016. bioRxiv 2021 (In eng). DOI: 10.1101/2021.02.17.431683.

18. CDC. COVID-19 Vaccinations in the United States. (https://covid.cdc.gov/covid=data-tracker/#vaccinations_vacc-total-admin-rate-total).

19. Brown CM, Vostok J, Johnson H, et al. Outbreak of SARS-CoV-2 Infections, Including COVID-19 Vaccine Breakthrough Infections, Associated with Large Public Gatherings - Barnstable County, Massachusetts, July 2021. MMWR Morb Mortal Wkly Rep 2021;70(31):1059–1062. (In eng). DOI: 10.15585/mmwr.mm7031e2.

20. Dougherty K, Mannell M, Naqvi O, Matson D, Stone J. SARS-CoV-2 B.1.617.2 (Delta) Variant COVID-19 Outbreak Associated with a Gymnastics Facility - Oklahoma, April-May 2021. MMWR Morb Mortal Wkly Rep 2021;70(28):1004–1007. (In eng). DOI: 10.15585/mmwr.mm7028e2.

21. Farinholt T, Doddapaneni H, Qin X, et al. Transmission event of SARS-CoV-2 delta variant reveals multiple vaccine breakthrough infections. BMC Med 2021;19(1):255. (In eng). DOI: 10.1186/s12916-021-02103-4.

22. Kang M, Xin H, Yuan J, et al. Transmission dynamics and epidemiological characteristics of Delta variant infections in China.medRxiv 2021:2021.08.12.21261991. DOI: 10.1101/2021.08.12.21261991.

23. Zhang M, Xiao J, Deng A, et al. Transmission Dynamics of an Outbreak of the COVID-19 Delta Variant B.1.617.2 - Guangdong Province, China, May-June 2021. China CDC Wkly 2021;3(27):584–586. (In eng). DOI: 10.46234/ccdcw2021.148.

24. Mallapaty S. Delta’s rise is fuelled by rampant spread from people who feel fine. Nature 2021 (In eng). DOI: 10.1038/d41586-021-02259-2.

25. Baden LR, El Sahly HM, Essink B, et al. Efficacy and Safety of the mRNA-1273 SARS-CoV-2 Vaccine. N Engl J Med 2021;384(5):403–416. (In eng). DOI: 10.1056/NEJMoa2035389.

26. Thomas SJ, Moreira ED, Kitchin N, et al. Safety and Efficacy of the BNT162b2 mRNA Covid-19 Vaccine through 6 Months. N Engl J Med 2021 (In eng). DOI: 10.1056/NEJMoa2110345.

27. CC-VBCI T. COVID-19 Vaccine Breakthrough Infections Reported to CDC - United States, January 1-April 30, 2021. MMWR Morb Mortal Wkly Rep2021:792–3.

28. Riemersma KK, Grogan BE, Kita-Yarbro A, et al. Shedding of Infectious SARS-CoV-2 Despite Vaccination when the Delta Variant is Prevalent - Wisconsin, July 2021. medRxiv 2021:2021.07.31.21261387. DOI: 10.1101/2021.07.31.21261387.

29. Singanayagam A, Patel M, Charlett A, et al. Duration of infectiousness and correlation with RT-PCR cycle threshold values in cases of COVID-19, England, January to May 2020. Euro Surveill 2020;25(32) (In eng). DOI: 10.2807/1560-7917.ES.2020.25.32.2001483.

30. Goyal A, Reeves DB, Cardozo-Ojeda EF, Schiffer JT, Mayer BT. Viral load and contact heterogeneity predict SARS-CoV-2 transmission and super-spreading events. Elife 2021;10 (In eng). DOI: 10.7554/eLife.63537.

31. Marks M, Millat-Martinez P, Ouchi D, et al. Transmission of COVID-19 in 282 clusters in Catalonia, Spain: a cohort study. Lancet Infect Dis 2021;21(5):629–636. (In eng). DOI: 10.1016/S1473-3099(20)30985-3.

32. Puranik A, Lenehan PJ, Silvert E, et al. Comparison of two highly-effective mRNA vaccines for COVID-19 during periods of Alpha and Delta variant prevalence. medRxiv 2021 (In eng). DOI: 10.1101/2021.08.06.21261707.

33. Altschul SF, Gish W, Miller W, Myers EW, Lipman DJ. Basic local alignment search tool. J Mol Biol 1990;215(3):403–10. (In eng). DOI: 10.1016/S0022-2836(05)80360-2.

34. Giovanetti M, Cella E, Benedetti F, et al. SARS-CoV-2 shifting transmission dynamics and hidden reservoirs potentially limit efficacy of public health interventions in Italy. Commun Biol 2021;4(1):489. (In eng). DOI: 10.1038/s42003-021-02025-0.

35. Moshiri N. ViralMSA: Massively scalable reference-guided multiple sequence alignment of viral genomes. Bioinformatics 2020 (In eng). DOI: 10.1093/bioinformatics/btaa743.

36. Minh BQ, Schmidt HA, Chernomor O, et al. IQ-TREE 2: New Models and Efficient Methods for Phylogenetic Inference in the Genomic Era. Mol Biol Evol 2020;37(5):1530–1534. (In eng). DOI: 10.1093/molbev/msaa015.

37. Turakhia Y, Thornlow B, Hinrichs AS, et al. Ultrafast Sample placement on Existing tRees (UShER) enables real-time phylogenetics for the SARS-CoV-2 pandemic. Nat Genet 2021;53(6):809–816. (In eng). DOI: 10.1038/s41588-021-00862-7.

38. Price MN, Dehal PS, Arkin AP. FastTree 2--approximately maximum-likelihood trees for large alignments. PLoS One 2010;5(3):e9490. (In eng). DOI: 10.1371/journal.pone.0009490.

39. Rambaut A, Lam TT, Max Carvalho L, Pybus OG. Exploring the temporal structure of heterochronous sequences using TempEst (formerly Path-O-Gen). Virus Evol 2016;2(1):vew007. DOI: 10.1093/ve/vew007.

40. Paradis E, Schliep K. ape 5.0: an environment for modern phylogenetics and evolutionary analyses in R. Bioinformatics 2019;35(3):526–528. (In eng). DOI: 10.1093/bioinformatics/bty633.

41. Prosperi MC, Salemi M. QuRe: software for viral quasispecies reconstruction from next-generation sequencing data. Bioinformatics 2012;28(1):132–3. DOI: 10.1093/bioinformatics/btr627.

42. Ragonnet-Cronin M, Hodcroft E, Hué S, et al. Automated analysis of phylogenetic clusters. BMC Bioinformatics 2013;14:317. (In eng). DOI: 10.1186/1471-2105-14-317.

43. Rai B, Shukla A, Dwivedi LK. Estimates of serial interval for COVID-19: A systematic review and meta-analysis. Clin Epidemiol Glob Health 2021;9:157–161. (In eng). DOI: 10.1016/j.cegh.2020.08.007.

44. CDC. COVID-19 Case Surveillance Public Use Data with Geography.

45. Grafen A. The phylogenetic regression. Philos Trans R Soc Lond B Biol Sci 1989;326(1233):119–57. (In eng). DOI: 10.1098/rstb.1989.0106.

46. Adams DC. A method for assessing phylogenetic least squares models for shape and other high-dimensional multivariate data. Evolution 2014;68(9):2675–88. (In eng). DOI: 10.1111/evo.12463.

47. Levine-Tiefenbrun M, Yelin I, Katz R, et al. Initial report of decreased SARS-CoV-2 viral load after inoculation with the BNT162b2 vaccine. Nat Med 2021;27(5):790–792. (In eng). DOI: 10.1038/s41591-021-01316-7.

48. Bar-On YM, Goldberg Y, Mandel M, et al. Protection of BNT162b2 Vaccine Booster against Covid-19 in Israel. N Engl J Med 2021;385(15):1393–1400. (In eng). DOI: 10.1056/NEJMoa2114255.

