## Supplemental Information for "SARS-CoV-2 Delta vaccine breakthrough transmissibility in Alachua, Florida"

**Table S1.** SARS-CoV-2 sequenced samples from different locales.

| | **Shands Hospital (Alachua)** [1] | | **University of Florida PathLabs (Alachua)** [2] | | **University of Miami (Miami-Dade)** [2] | | **BayCare (Tampa Bay)** [2] | |
|---|---|---|---|---|---|---|---|---|
| ***Year/month*** | *Total* | *High coverage* [3] | *Total* | *High coverage* [3] | *Total* | *High coverage* [3] | *Total* | *High coverage* [3] |
| 2020-10 | 5 | 4 | 0 | 0 | 12 | 0 | 0 | 0 |
| 2020-11 | 90 | 68 | 0 | 0 | 7 | 0 | 0 | 0 |
| 2020-12 | 503 | 389 | 0 | 0 | 0 | 0 | 0 | 0 |
| 2021-01 | 589 | 417 | 346 | 242 | 0 | 0 | 15 | 14 |
| 2021-02 | 253 | 145 | 555 | 347 | 138 | 78 | 11 | 9 |
| 2021-03 | 174 | 78 | 207 | 139 | 460 | 318 | 5 | 4 |
| 2021-04 | 176 | 125 | 98 | 63 | 349 | 240 | 33 | 25 |
| 2021-05 | 196 | 115 | 5 | 4 | 158 | 97 | 46 | 31 |
| 2021-06 | 83 | 43 | 4 | 4 | 0 | 0 | 44 | 42 |
| 2021-07 | 114 | 91 | 850 | 673 | 93 | 77 | 0 | 0 |
| 2021-08 | 25 | 22 | 166 | 141 | 177 | 144 | 0 | 0 |
| Total | 2,208 | 1,497 (68%) | 2,231 | 1,613 (72%) | 1,394 | 954 (68%) | 154 | 125 (81%) |

1. Samples from NP swab
2. Samples from saliva
3. >70% coverage, at least 20X depth

**Table S2**. Adjusted and unadjusted linear regression estimates for the relationships between patient characteristics and level of infectivity among COVID-19 vaccine breakthrough cases in Alachua County, Florida, from February – July, 2021.

| | **Viral load level (RNA Log copies/ml)** | | | |
|---|---|---|---|---|
| | **Unadjusted** | | **Adjusted** | |
| | **Estimate** | **Pr(>\|t\|)** | **Estimate** | **Pr(>\|t\|)** |
| **Age** (continuous years) | 0.008641 | 0.618 | -- | -- |
| **Age (**categorical years) | | | | |
| 30-49 *vs. 18-29* | 1.0208 | 0.334 | 0.924903 | 0.394 |
| 50+ *vs. 18-29* | -1.8006 | 0.139 | -1.901733 | 0.134 |
| **Sex** | | | | |
| Male *vs. female* | -0.5399 | 0.265 | -- | -- |
| **Race** | | | | |
| White *vs. non-white* | 0.7012 | 0.202 | -- | -- |
| **Ethnicity** | | | | |
| Hispanic *vs. Non- Hispanic* | 0.1277 | 0.83 | -- | -- |
| **Vaccine** | | | | |
| Moderna *vs. J&J* | 0.8256 | 0.437 | -1.528243 | 0.472 |
| Pfizer *vs. J&J* | 0.9049 | 0.257 | -1.786090 | 0.265 |
| **Variant** | | | | |
| Delta vs. *non-Delta or unknown* | 0.3645 | 0.45 | -0.891119 | 0.390 |
| **Time between vaccination and onset** (continuous days) | -0.0003594 | 0.931 | 0.007665 | 0.400 |
| **Time between vaccination and onset** (categorized) | | | | |
| >3 months *vs. 0-3 months* | 0.02763 | 0.954 | -- | -- |
| **Time between onset and sample collection** (continuous days) | -0.009568 | 0.927 | -- | -- |

* Indicates variable was not considered in adjusted analysis

**Table S3**. Adjusted and unadjusted logistic regression estimates for the relationships between patient characteristics and level of infectivity among COVID-19 vaccine breakthrough cases in Alachua County, Florida, from February – July, 2021.

| | **Viral load level (RNA Log copies/ml) ≥ Log 4 *vs.* < Log 4** | | | |
|---|---|---|---|---|
| | **Unadjusted ORs** | **P-value** | **Adjusted ORs** | **P-value** |
| **Age** (continuous years) | 1.02 (0.98-1.05) | 0.336 | -- | |
| **Age (**categorical years) | | | | |
| 30-49 *vs. 18-29* | 0.78 (0.30-2.01) | 0.611 | 0.82 (0.30-2.22) | 0.6970 |
| 50+ *vs. 18-29* | 2.24 (0.75-7.14) | 0.159 | 2.39 (0.75-8.22) | 0.1501 |
| **Sex** | | | | |
| Male *vs. female* | 0.65 (0.27-1.52) | 0.321 | -- | -- |
| **Race** | | | | |
| White *vs. non-white* | 1.31 (0.50-3.49) | 0.584 | -- | -- |
| **Ethnicity** | | | | |
| Hispanic *vs. Non- Hispanic* | 1.70 (0.59-4.98) | 0.564 | -- | -- |
| **Vaccine** | | | | |
| Moderna *vs. J&J* | 1.56 (0.24-10.72) | 0.638 | 1.77 (0.24-14.26) | 0.5757 |
| Pfizer *vs. J&J* | 1.56 (0.24-10.72) | 0.783 | 1.59 (0.34-8.07) | 0.5574 |
| **Variant** | | | | |
| Delta vs. *non-Delta or unknown* | 2.46 (1.05-5.97) | 0.0414 | 3.04 (1.16-8.54) | 0.0274 |
| **Time between vaccination and onset** (continuous days) | 1.00 (0.99-1.01) | 0.748 | 0.99 (0.99-1.00) | 0.1602 |
| **Time between vaccination and onset** (categorized) | | | | |
| >3 months *vs. 0-3 months* | 0.96 (0.41-2.24) | 0.932 | -- | -- |
| **Time between onset and sample collection** (continuous days) | 0.98 (0.81-1.17) | 0.817 | -- | -- |

* Indicates variable was not considered in adjusted analysis

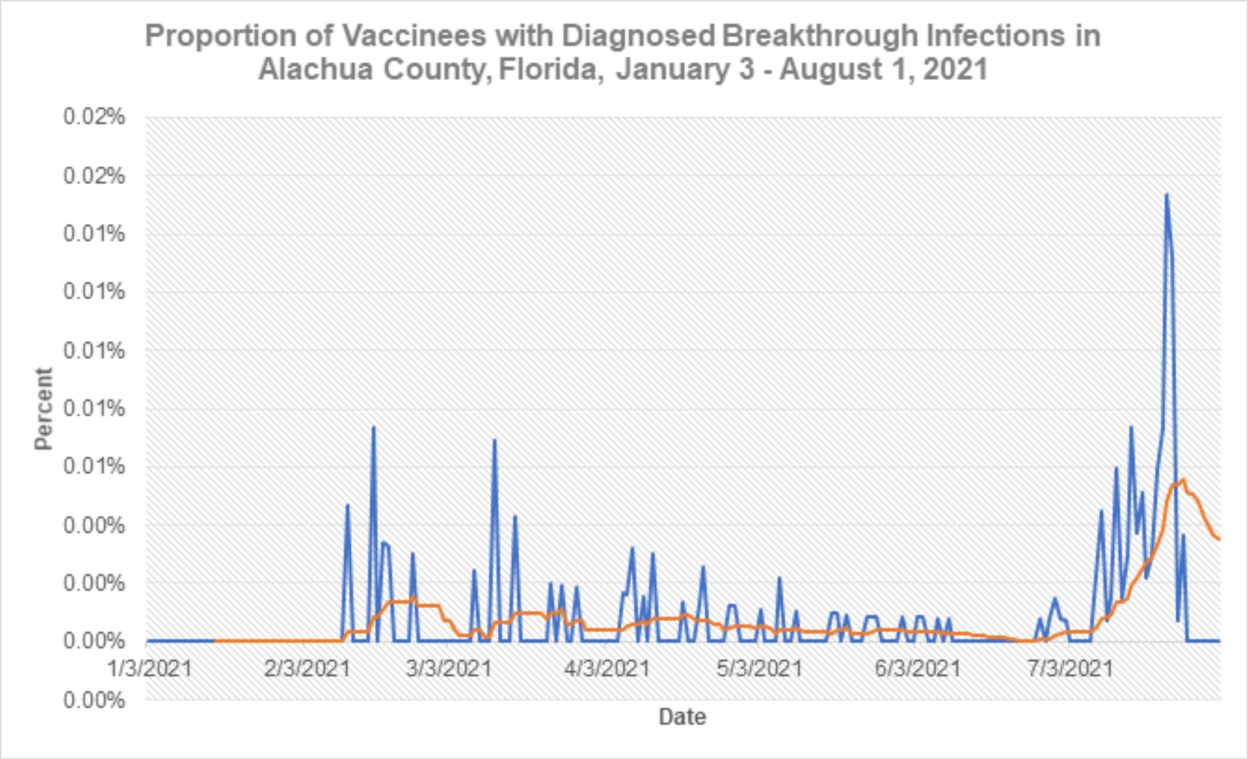


**Figure S1. Number (blue) of vaccinations and proportion with diagnosed breakthrough infections (orange) in Alachua County, Florida, from January 03 - August 01, 2021.**

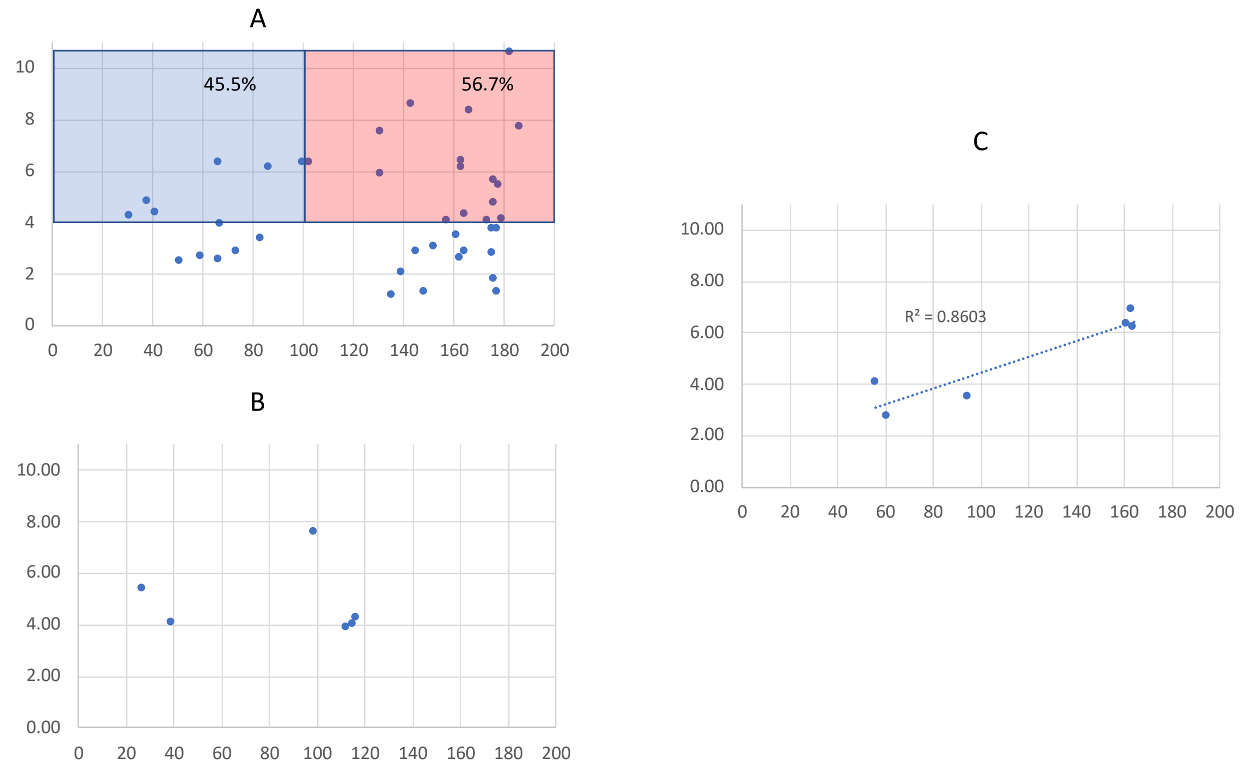


**Figure S2**. **VL vs. time from full vaccination at the time of symptoms onset in vaccine breakthrough cases infected with the Delta variant by vaccine type.** Each dot in the scatterplots represents a single patient fully vaccinated with either Pfizer/BNT162b2 (Panel A), Johnson & Johnson/Janssen (Panel B), or Moderna/mRNA-1273 (Panel C) vaccine. In each graph, The Y-axis reports the VL measure in Log RNA copies/ml. The x-axis represents the time (in days) between full vaccination, defined as two weeks after 2$^{nd}$ vaccination dose (in case of Pfizer-BioNTech or Moderna) or after single dose (Johnson & Johnson/Janssen), and time of sampling, which occurred for each patient on average 4.2 days after symptoms onset (see Table 1). Shaded areas in (A) highlight proportions of patients with VL above the transmissibility threshold (Log 4 copies/ml) who were fully vaccinated for <101 days (cyan) or >100 (pink).
